# Fast Evaluation of Viral Emerging Risks (FEVER): A computational tool for biosurveillance, diagnostics, and mutation typing of emerging viral pathogens

**DOI:** 10.1101/2021.05.25.21257811

**Authors:** Zachary R. Stromberg, James Theiler, Brian T. Foley, Adán Myers y Gutiérrez, Attelia Hollander, Samantha J. Courtney, Jason Gans, Alina Deshpande, Ebany J. Martinez-Finley, Jason Mitchell, Harshini Mukundan, Karina Yusim, Jessica Z. Kubicek-Sutherland

## Abstract

Viral pathogen can rapidly evolve, adapt to novel hosts and evade human immunity. The early detection of emerging viral pathogens through biosurveillance coupled with rapid and accurate diagnostics are required to mitigate global pandemics. However, RNA viruses can mutate rapidly, hampering biosurveillance and diagnostic efforts. Here, we present a novel computational approach called FEVER (Fast Evaluation of Viral Emerging Risks) to design assays that simultaneously accomplish: 1) broad-coverage biosurveillance of an entire class of viruses, 2) accurate diagnosis of an outbreak strain, and 3) mutation typing to detect variants of public health importance. We demonstrate the application of FEVER to generate assays to simultaneously 1) detect sarbecoviruses for biosurveillance; 2) diagnose infections specifically caused by severe acute respiratory syndrome coronavirus 2 (SARS-CoV-2); and 3) perform rapid mutation typing of the D614G SARS-CoV-2 spike variant associated with increased pathogen transmissibility. These FEVER assays had a high *in silico* recall (predicted positive) up to 99.7% of 525,708 SARS-CoV-2 sequences analyzed and displayed sensitivities and specificities as high as 92.4% and 100% respectively when validated in 100 clinical samples. The D614G SARS-CoV-2 spike mutation PCR test was able to identify the single nucleotide identity at position 23,403 in the viral genome of 96.6% SARS-CoV-2 positive samples without the need for sequencing. This study demonstrates the utility of FEVER to design assays for biosurveillance, diagnostics, and mutation typing to rapidly detect, track, and mitigate future outbreaks and pandemics caused by emerging viruses.

## INTRODUCTION

In the last decade, several RNA viruses have emerged from animal reservoirs to threaten global public health (1). These viruses include influenza A in 2009 (2), Ebola in 2013 (3), Chikungunya in 2014 (4), Zika in 2015 (5), and severe acute respiratory syndrome coronavirus 2 (SARS-CoV-2) most recently in 2019 (6, 7). Outbreaks and pandemics often occur when a zoonotic viral strain evolves and escapes human immunity (8, 9). The early detection of these pathogens is required to minimize outbreaks and prevent pandemics by guiding early interventions (10). However, RNA viruses mutate rapidly, so biosurveillance tools must be able to detect groups of viral pathogens with diverse genome sequences and emerging variants, often requiring time-consuming and expensive procedures performed by trained personnel that are not easily accessible in all regions of the world (11, 12). Once spillover occurs and a viral isolate enters human circulation, a rapid and accurate diagnostic test is required to circumvent its spread (13-16). However, development of accurate diagnostics at the onset of an outbreak can be challenging due to limited data availability depending on the location of the outbreak, and the process for validation and implementation of a new test can also be time-consuming (17). Having a broadly applicable computational approach to assay design that can accommodate pathogen emergence and variant evolution would help alleviate some of these challenges.

Coronaviruses are positive-sense, single-stranded RNA viruses and a leading cause of the common cold (18). However, several recent coronavirus outbreaks, including SARS-CoV in 2002-2003, and Middle East respiratory syndrome coronavirus (MERS-CoV) in 2012, were associated with significant morbidity and mortality (8, 19, 20). In December 2019, SARS-CoV-2 emerged in humans and spread rapidly, resulting in the COVID-19 (coronavirus disease 2019) global pandemic (21-23). SARS-CoV-2, along with SARS-CoV and SARS-like viruses, belong to the subgenus *Sarbecovirus* (24). Effective diagnostic and biosurveillance assays are critical to control the spread of SARS-CoV-2 and prevent future *Sarbecovirus* pandemics.

Real-time reverse transcription PCR (RT-PCR) is considered the gold standard for COVID-19 diagnostics (25) with assays developed by the United States Centers for Disease Control and Prevention (U.S. CDC) (26), China CDC (27), Hong Kong University (28), and Charité Institute of Virology (29), along with commercial products (30). Traditionally, RT-PCR probe and primer design involved selecting a target from a specific gene, often a virulence factor (31). In contrast, modern probe and primer design involves multiple sequence alignments and algorithms to determine conserved signatures for detection (32-34). However, if assays are not updated periodically to account for variants, genome mutations can emerge at the primer or probe binding site and limit assay utility (35, 36). The U.S. CDC initially designed a pan-*Sarbecovirus* assay targeting the nucleocapsid gene (2019-nCoV_N3) with broad coverage, however it eliminated this assay from its diagnostic panel to simplify the testing process (37) and because of cross-reactivity (38) and false-negative results on clinical samples (39). Also, mutations at either a primer or probe binding site (not specified because of proprietary components) yielded false-negative results for the Roche cobas envelope gene pan-*Sarbecovirus* assay (36). Finally, mutations have made SARS-CoV-2 more transmissible (40, 41) and potentially more resistant to immunity induced by vaccination (42). The current practice of independently introducing biosurveillance efforts, targeted diagnostics and mutation tracking has hampered our ability to adapt and respond to the dynamic challenges presented by the COVID-19 pandemic. The development of a synchronous approach to integrate these data streams can help mitigate the challenges posed by emerging viral pathogens and support rapid decision making to help combat future outbreaks and pandemics.

There is a critical need for tools that can be used in biosurveillance and diagnostic applications to respond rapidly to emerging pathogens and mitigate their global impact (43). To this end, we have developed FEVER (Fast Evaluation of Viral Emerging Risks), a computational approach that can generate both high-coverage or strain-specific diagnostic assays so that the same detection platform can be used for both broad-based biosurveillance and targeted diagnostic applications. Here, we demonstrate the applicability of FEVER for simultaneous biosurveillance of pan-*Sarbecovirus*es and targeted diagnosis of SARS-CoV-2 in a cohort of 100 human patients. We also demonstrate mutation typing of the D614G SARS-CoV-2 spike variant using this approach as an example of a rapid method for tailored surveillance of pathogen evolution and emergence.

## MATERIALS AND METHODS

### Ethics

This study was designed in alignment with DOE, NIH, and universal HIPAA guidelines. The protocol was reviewed and approved by the Institutional Review Board of the Los Alamos National Laboratory (LANL#000473), per DOE guidelines and policies, and by the Presbyterian Institutional Review Board (PHS IRB#1608836).

### FEVER surveillance and diagnostic probe and primer design

For the surveillance and diagnostics FEVER assays, four sets of primers and probes were generated (Table 1). Two sets of primers and probes were developed for universal detection of all sarbecoviruses, with one targeting the 5’ untranslated region (UTR) termed FEVER_5’UTR and another targeting the envelope gene termed FEVER_Env. Another two sets of primers and probes were developed for specific detection of SARS-CoV-2, with one targeting the ORF1ab region termed FEVER_ORF1ab and one targeting the spike gene called FEVER_Spike. After the FEVER probes were designed, primers flanking the probes were created by the PrimerDesign-M tool (44, 45) using an alignment of SARS-CoV-2 sequences.

**Table 1.**
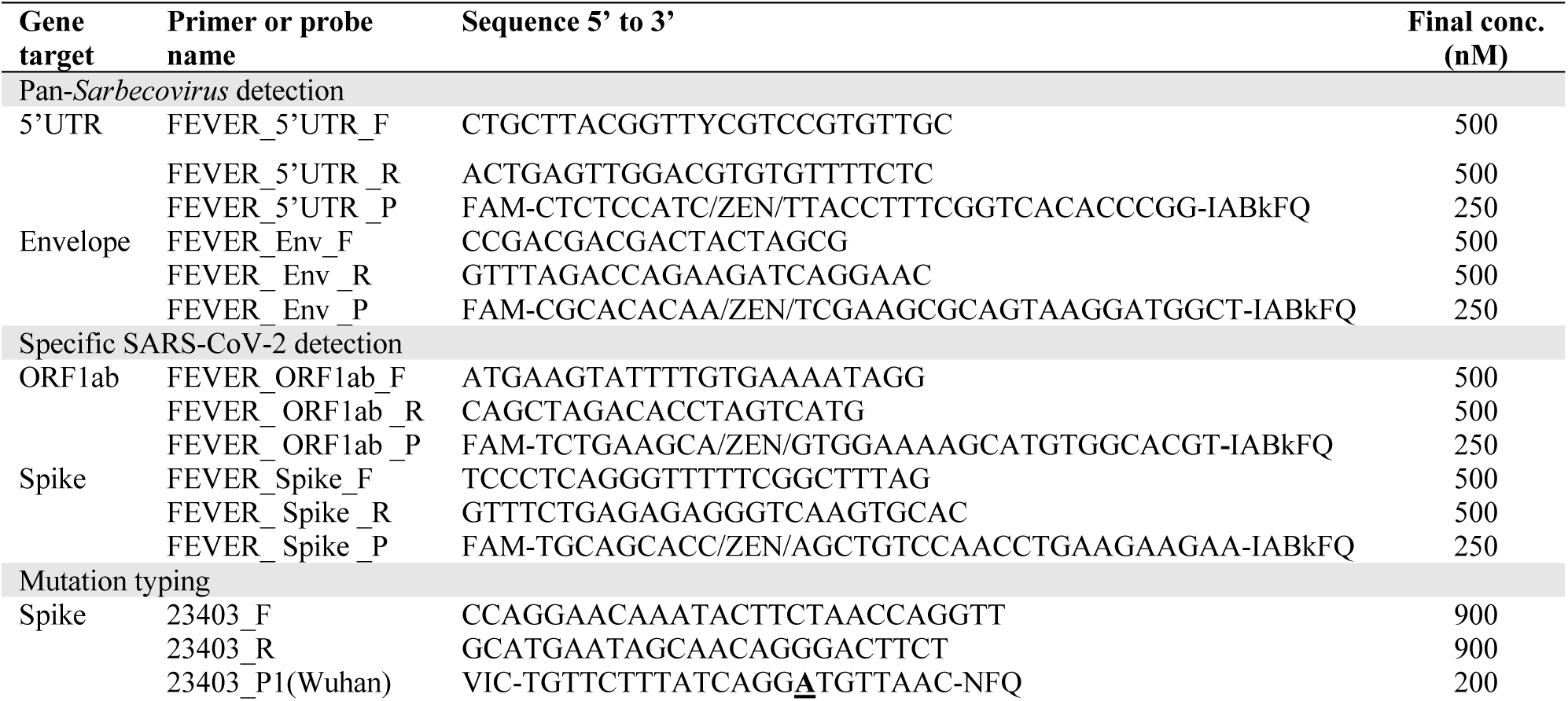
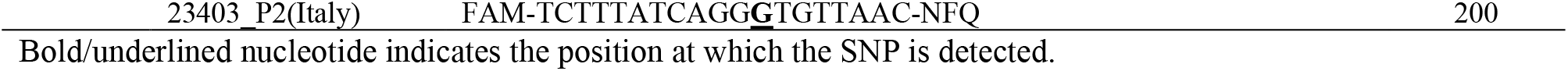
FEVER Assays for COVID-19.

#### Sequence data

As of March 10, 2020 the Global Initiative on Sharing All Influenza Data (GISAID) database (https://www.epicov.org or https://www.gisaid.org/) had acquired 338 complete, or near-complete, high-quality SARS-CoV-2 genome sequences. After aligning the sequences and eliminating all but one of each set of sequences which were 100% identical to each other to reduce redundancy, our data set consisted of 19 unique sequences of SARS-CoV-2. We also collected all coronaviruses from GenBank that were in the “Sarbecovirus clade” of the beta coronaviruses, using BLAST searches rather than GenBank taxonomy because many sarbecoviruses were listed as “unclassified coronavirus” in the taxonomy table. Again, we reduced redundancy be eliminating identical sequences (primarily many dozens of SARS sequences from the 2003-2004 outbreak), and retained a set of 67 genome sequences. It was these 86 *Sarbecovirus* genomes that were used to design our assays.

#### FEVER algorithm

The FEVER algorithm does not require sequence alignments prior to probe design. Instead, our probe design goal was to produce a short nucleotide sequence (called a “*k*-mer”, *e*.*g*., a short DNA sequence, *k* base pairs long) that binds to SARS-CoV-2 without binding to background material that may be present in a sample that is being tested. Specific desirable properties of the probe included maintaining a high GC-content and reducing the hairpin propensity (*i*.*e*., the tendency of the probe string to fold over on itself) by ensuring that any self-complementary pairs of sub-strings in the probe are sufficiently short. The mathematical formalization of the problem was expressed in terms of a constrained optimization, with minimum GC-content and the maximum length of self-complementary sub-strings taken as constraints while maximizing coverage. A probe is said to “cover” a SARS-CoV-2 sequence if the probe’s *k*-mer appears as a sub-sequence of that sequence. Thus, coverage is defined as the number of sequences in the database covered by the probe. “Exact” coverage corresponds to an exact match of probe and sub-sequence. Hamming distance between two strings is defined as the number of characters that would have to be changed in one string to agree with the other string. Given a database of SARS-CoV-2 sequences, the single probe design problem is to find a string of length *k*, subject to the constraints of GC-content and hairpin aversion, that covers as many SARS-CoV-2 sequences as possible. For the multi-probe design problem, we seek *n* probes instead of just one, and a *n*-probe design is said to cover a SARS-CoV-2 sequence if any one of the probes covers it. The selection of *k*=31 was used because of practical considerations for molecular beacon synthesis allowing for a 12 bp stem region (6 bp on each side of the *k*-mer). Also, *k*=31 was long enough to bind to target sequences in the SARS-CoV-2 database without accidentally covering potential background sequences.

Our FEVER algorithm works by extracting all *k*-mers from each of the sequences in the database and then restricting consideration to those that exhibit a minimal GC-content and hairpin aversion. For every one of these *k*-mers, we counted how many viral sequences were covered, being careful not to count a sequence twice even if the *k*-mer appears twice in it. Finally, we selected the *k*-mer with the largest count. To ensure that our probes were not susceptible to viral mutation, a multi-probe design approach was applied. The first probe was designed using a naïve approach by producing the optimal probe with highest coverage of the queried sequence database. The next probe is designed by eliminating the viral sequences covered by the first probe and using this truncated database to find the single probe that best covers the remaining viral sequences. This process is repeated until 100% of the database is covered. For both pan-*Sarbecovirus* sequences and SARS-CoV-2 sequences it only took 2 probes to obtain 100% coverage of the sequence database at the time of design.

#### Data availability

All code written in support of this publication is publicly available at https://github.com/jt-lanl/fever-probes.

### FEVER mutation typing probe and primer design

A variant carrying the amino acid change D614G spike mutation became globally dominant, and the G614 variant has been demonstrated to have a fitness advantage associated with higher viral loads (40). A custom TaqMan SNP Genotyping Assay was designed (Thermo Fisher, 4332075) to detect this single nucleotide polymorphism (SNP) in SARS-CoV-2 using Thermo Fisher Scientific’s Custom Assay Design Tool. Probe and primer sequences are presented in Table 1.

### Analysis of primer and probe coverage

The FEVER biosurveillance and diagnostics assays were computationally characterized using two complementary methods. First, the frequency of SARS-CoV-2 variants in the amplified region of the FEVER and U.S. CDC assays were assessed visually using the Variant Visualizer, which is under development and will be available at https://cov.lanl.gov. Second, we used an *in silico* validation tool (46) to assess the inclusivity of the FEVER assays compared with the U.S. CDC N1 and N2 assays (26). Each assay (forward primer, reverse primer and TaqMan probe) was evaluated against each SARS-CoV-2 sequence from GISAID (47) public database that was greater than 29 kb (*n* = 525,708) to ensure only complete genomes were assessed. The GISAID sequence data used in this analysis was accessed on February 15, 2021. Results are presented in Table 2. For this analysis, a false-negative was assigned if one or more assay oligonucleotides satisfied any of the following conditions: (i) three or more mismatches to a target sequence, (ii) a predicted melting temperature less than 40°C between the oligonucleotide and a target sequence, or (iii) when primer/target mismatches occurred in either of the last two 3’ bases of a primer associated with an expected increase of 2 or more in the cycle threshold (C_T_) value (as defined previously (48)). True-positives were assigned for any assay/target sequence pairing that did *not* result in a false negative. Results are also reported as the number of genome sequences that contained perfect matches, single mismatches, two mismatches, or three or more mismatches for a given assay.

**Table 2.** *In silico* inclusivity test against SARS-CoV-2 sequences.

| Assay | Recall (%)<sup>a</sup> | No. sequences with perfect match | No. sequences with 1 mismatch | No. sequences with 2 mismatches | No. sequences that failed<sup>b</sup> |
| --- | --- | --- | --- | --- | --- |
| FEVER_5'UTR | 96.39 | 413,603 | 91,763 | 1,361 | 18,981 |
| FEVER_Env | 98.63 | 511,976 | 6,164 | 343 | 7,225 |
| FEVER_ORF1ab | 99.72 | 490,388 | 33,723 | 143 | 1,454 |
| FEVER_Spike | 90.51 | 360,033 | 115,321 | 481 | 49,873 |
| U.S. CDC N1 | 98.95 | 506,092 | 11,454 | 2,633 | 5,529 |
| U.S. CDC N2 | 99.17 | 507,298 | 13,901 | 156 | 4,353 |

Oligonucleotides from each assay were assessed against 525,708 sequences of SARS-CoV-2 obtained from GISAID on February 15, 2021.
<sup>a</sup>Recall = true-positive (sum of the perfect match, single mismatch, and double mismatch) / (true-positive + false negative).
<sup>b</sup>Failure represents SARS-CoV-2 sequences that were not detected (false-negative).

### Patient sample collection

From November 2020 to December 2020, 100 nasopharyngeal (NP) swabs were collected from individuals suspected of having COVID-19 disease or otherwise randomly tested in the U.S. state of New Mexico. NP swabs were collected using existing U.S. CDC guidelines, labeled with a de-identified bar code, and stored under refrigerated conditions until batch shipping (within 24 hours of sample collection). Specimens were packed and shipped according to U.S. Department of Transportation regulations regarding the shipment of biological substances.

### RNA extraction

For spiked samples, RNA extraction was performed using the QIAamp Viral RNA Mini Kit (Qiagen). For the NP swab samples, RNA was extracted using the MagMAX Viral/Pathogen Nucleic Acid Isolation Kit (Thermo Fisher) and a KingFisher. For both kits, a 200 µL portion of each sample was used for extraction following the manufacturer’s instructions with a 50 µL elution, and 5 µL of the eluted template was used in RT-PCR.

### RT-PCR protocol

Samples were tested using our FEVER and U.S. CDC 2019-novel coronavirus (2019-nCoV) RT-PCR diagnostic panel (termed U.S. CDC assays) (26). For the U.S. CDC N1 (2019-nCoV_N1) and N2 (2019-nCoV_N2) assays, each 20 µL reaction consisted of 8.5 µL of water, 1.5 µL of combined primer and probe mix (IDT, 500 nM primer and 125 nM probe, final concentrations), 5 µL of TaqPath 1-step RT-qPCR master mix (Thermo Fisher), and 5 µL of the template. For the FEVER assays (FEVER_5’UTR, FEVER_Env, FEVER_ORF1ab, and FEVER_Spike), each 20 µL reaction consisted of 8 µL of water, 2 µL of combined primer and probe mix (IDT, 500 nM primer and 250 nM probe, final concentrations), 5 µL of TaqPath 1-step RT-qPCR master mix, and 5 µL of the template. The thermocycling conditions were the same for all assays. Thermocycling conditions using an ABI StepOnePlus or 7500 Fast Dx were 25°C for 2 min, 50°C for 15 min, 95°C for 2 min, followed by 45 cycles of 95°C for 3 s and 55°C for 30 s. For RT-PCR assays, samples were considered positive when the C_T_ value ≤40. Patient clinical samples were considered positive for SARS-CoV-2 when the two U.S. CDC assays N1 and N2 both had a C_T_ value ≤40, and negative when both targets had a C_T_ value >40 according to CDC guidelines (37). If only one N1 or N2 result was positive, the result was considered inconclusive and the sample was re-run. If the re-test results were also inconclusive, the final result was reported as inconclusive.

### Analytical specificity of RT-PCR

The analytical specificities of the FEVER and U.S. CDC assays were evaluated using genomic RNA from various viruses obtained from BEI resources. We tested 5 µL of undiluted genomic RNA preparation from SARS-CoV Urbani (NR-52346), avian coronavirus Massachusetts (NR-49096), alphacoronavirus Purdue P115 (NR-48571), porcine respiratory coronavirus ISU-1 (NR-48572), two influenza A viruses (NR-20080 and NR-20081), and two influenza B viruses (NR-45848 and NR-45849). RNA from heat-inactivated SARS-CoV-2 isolate USA-WA1/2020 (BEI Resources, NR-52347) was a positive assay control, and water was a negative assay control.

### Analytical sensitivity

The analytical sensitivity of each assay was determined by spiking inactivated SARS-CoV-2 USA-WA1/2020 strain (BEI resources, NR-52286) at 10^4^ copies/µL, and 10-fold serial dilutions were performed in an NP swab matrix. NP swabs from a single human donor (Lee Biosolutions, #991-31-NC-COVID-N) were suspended in 0.45% saline and confirmed SARS-CoV-2 negative by the supplier. Each concentration was tested using three biological replicates.

### Assessment of NP swab samples

The 100 NP swab samples were transported to the laboratory for RNA extraction and testing. After RNA extraction, samples were stored at –80°C. Samples were tested by our FEVER and U.S. CDC assays for SARS-CoV-2. For the U.S. CDC assays, the 2019-Ncov plasmid (IDT) containing the nucleocapsid gene was used as a positive control, and water was used as a negative control. For the FEVER assays, RNA from heat-inactivated SARS-CoV-2, isolate USA-WA1/2020 (BEI Resources, NR-52347) was used as a positive control, and water was used as a negative control. The RNase P assay described by the U.S. CDC assays (26) was used as an extraction control for each sample.

### Mutation typing of SARS-CoV-2 by PCR

Extracted RNA (10 µL) from SARS-CoV-2 positive patient samples was reverse transcribed in a 20 µL reaction using the High Capacity cDNA Reverse Transcription Kit (Thermo Fisher, 4368814) according to the manufacturer’s instructions. Each 20 µL SNP genotyping PCR reaction consisted of 5 µL of cDNA as the template, 4.5 µL of water, 0.5 µL of 40× SNP genotyping mix (900 nM primer and 200 nM probe final concentrations), and 10 µL of 2× TaqMan Genotyping Master Mix (Thermo Fisher, 4371353). Amplification was performed using an ABI StepOnePlus RT-PCR machine under the following conditions: 10 min at 95°C and 45 cycles of 15 s at 95°C and 60 s at 60°C in a 96-well plate. The PCR C_T_ values corresponding with the wild type (VIC) and mutant (FAM) forms were compared, and the lower value (≤45) of the two was determined to be the SNP sequence. Samples were considered undetermined when both reactions had a C_T_ value >45. Control RNA from Heat-Inactivated SARS-CoV-2, isolate USA-WA1/2020 (BEI Resources, NR-52347) was used as a positive control, diluted to 10^4^ copies/µL, and run with each batch of cDNA synthesis and PCR. Isolate USA-WA1/2020 contains the wild-type spike sequence, and all results with this control showed that the wild-type C_T_ was lower than the mutant C_T_ value (data not shown).

A total of 17 NP swab samples had sufficient sample volume to be used in sequencing to confirm the D614G mutation. The spike gene was amplified using the 23403F_seq and 23403R_seq primers (5’-TGAACTTCTACATGCACCAGC −3’ and 5’-AAACAGCCTGCACGTGTTTG −3’, respectively). Each 25 µL reaction consisted of 5 µL of cDNA as the template, 11.75 µL of water, 5 µL of 5X Q5 reaction buffer (New England BioLabs), 0.5 µL of 10 mM dNTPs, 1.25 µL of each primer (0.5 µM final concentration), and 0.25 µL of Q5 hot start high-fidelity DNA polymerase (New England BioLabs). PCR was performed on a thermocycler as follows: 30 s at 95°C and 35 cycles of 10 s at 98°C, 30 s at 65°C, and 30 s at 72°C. Amplified products were purified using the PureLink PCR Purification Kit (Thermo Fisher, K310002) and were sent for sequencing to Eurofins Genomics.

### Statistical analysis

Data were analyzed with GraphPad Prism version 8. Confidence intervals were calculated using the modified Wald method. The Pearson correlation coefficient was used to assess the concordance between PCR C_T_ values. *P* values < 0.05 were deemed significant.

## RESULTS

### FEVER Computational Tool

FEVER combines probes and primers generated to 1) broadly identify an entire class of virus; 2) specifically detect a strain of interest; and 3) detect SNPs of importance to public health. The computational approach starts with a curated multiple sequence alignment. The FEVER algorithm does not however require sequences to be aligned in order to generate assays. The alignment was performed to avoid overrepresentation of any single isolate and ensure broad coverage of the pathogen of interest. In March of 2020, we developed two alignments using the sequences available in GISAID: one with SARS-CoV-2 sequences and one with *Sarbecovirus* sequences. Of note our FEVER assays were developed within one month of the U.S. CDC assays (41) and also used viral sequence information obtained from GenBank. Next, our FEVER algorithm was applied to design probes using the *Sarbecovirus* alignment for biosurveillance and the SARS-CoV-2 alignment for diagnostics while also screening the results using parameters set for GC-content, hairpin propensity, and match recall to achieve high coverage as well as reagent manufacturing and performance compatibility. The FEVER algorithm identifies sets of probes that cover 100% of each multiple sequence alignment. Two pan-*Sarbecoviruses* (FEVER_5’UTR and FEVER_Env) and two SARS-CoV-2 (FEVER_ORF1ab and FEVER_Spike) specific probes were needed to detect all input sequences. After the FEVER probes were designed, primers flanking the probe sites were designed using the PrimerDesign-M tool (44, 45) for use in RT-PCR assays (Table 1). Few variants were visually detected in the region of amplification of these primers and probes (Fig. S1). Further, we designed TaqMan SNP Genotyping PCR probes and primers to detect the previously identified A23403G (D614G amino acid change) mutation in the spike gene (40) (Table 1). Mutations in the SARS-CoV-2 genome conferring a fitness advantage have been reported (40). However, rapid methods to characterize SARS-CoV-2 mutations are lacking. Together, the primers and probes for all three components (biosurveillance, diagnostics, and mutation typing) are referred to collectively as the FEVER assays.

### *In silico* inclusivity test

We evaluated our FEVER assays using a public web-based validation tool against 525,708 sequences of SARS-CoV-2 (46). The *in silico* evaluation only included complete SARS-CoV-2 genome sequences that were at least 29 kb long. In general, a low number of predicted failures (false-negatives) was observed in comparison with the number of successes or true-positives (total of perfect matches, single mismatch, and double mismatches) (Table 2). Recall (true positives divided by the sum of true positives and false negatives) was used to assess relative assay performance. Of the FEVER and U.S. CDC assays, the SARS-CoV-2 specific FEVER probe targeting ORF1ab had the best-predicted performance with a recall of 99.7%. The pan-*Sarbecovirus* probe targeting the 5’ UTR and envelope gene were also predicted to be highly inclusive for SARS-CoV-2 with recalls of 96.4% and 98.6%, respectively. In contrast, the SARS-CoV-2 specific probe targeting the spike gene had a 90.5% recall, suggesting that the spike gene is a suboptimal target due to its high genetic variability (40, 49). For instance, the U.S. CDC N1 assay had a recall of 99.0% and the U.S. CDC N2 assay had a 99.2% recall. Compared with the U.S. CDC assays, the FEVER_ORF1ab assay had a ∼1% higher recall, FEVER_Env had a similar recall, and the FEVER_5’UTR and FEVER_Spike assays had lower recall values.

### Analytical specificity

Cross-reactivity of primers and probes with other microorganisms can reduce the efficacy of RT-PCR (50). For analysis of specificity, SARS-CoV-2 was the only target virus for the SARS-CoV-2 specific assays, and all sarbecoviruses were considered the targets for the pan-*Sarbecovirus* probes. We initially evaluated cross-reactivity *in silico* and then experimentally with non-target viruses. The specificity of each primer and probe set was tested *in silico* using BLAST, which yielded matches to SARS-CoV-2 only for the assay oligonucleotides specific to SARS-CoV-2 (FEVER_ORF1ab and FEVER_Spike) and matches to sarbecoviruses for the pan-*Sarbecovirus* assay oligonucleotides (FEVER_5’UTR and FEVER_Env). No hits to human DNA or other respiratory viruses were found.

For experimental evaluation of specificity, the FEVER assays were tested against three animal coronavirus strains (alphacoronavirus 1 strain Purdue P115, avian coronavirus strain Massachusetts, and porcine respiratory coronavirus strain ISU-1), four influenza virus strains (A/Brisbane/59/2007, A/Brisbane/10/2007, B/Nevada/03/2011, and B/Texas/06/2011), and a single strain of SARS-CoV (Urbani, isolated in 2003). The SARS-CoV-2 strain USA-WA1/2020 was used as a positive control. The three animal coronavirus and four influenza virus strains all tested negative (Table 3). Excluding SARS-CoV-2, the SARS-CoV Urbani strain was the only commercially available genomic RNA from a *Sarbecovirus* available at the time of testing. The SARS-CoV Urbani strain was correctly tested negative by the U.S. CDC assays (N1 and N2), FEVER_ORF1ab, and FEVER_Spike targets. The SARS-CoV strain Urbani tested positive by the pan-*Sarbecovirus* (FEVER_5’UTR and FEVER_Env) FEVER assays, which was expected because these were designed to detect all sarbecoviruses. The SARS-CoV-2 strain USA-WA1/2020 used as a positive control was correctly identified by both FEVER and U.S. CDC assays.

**Table 3.**
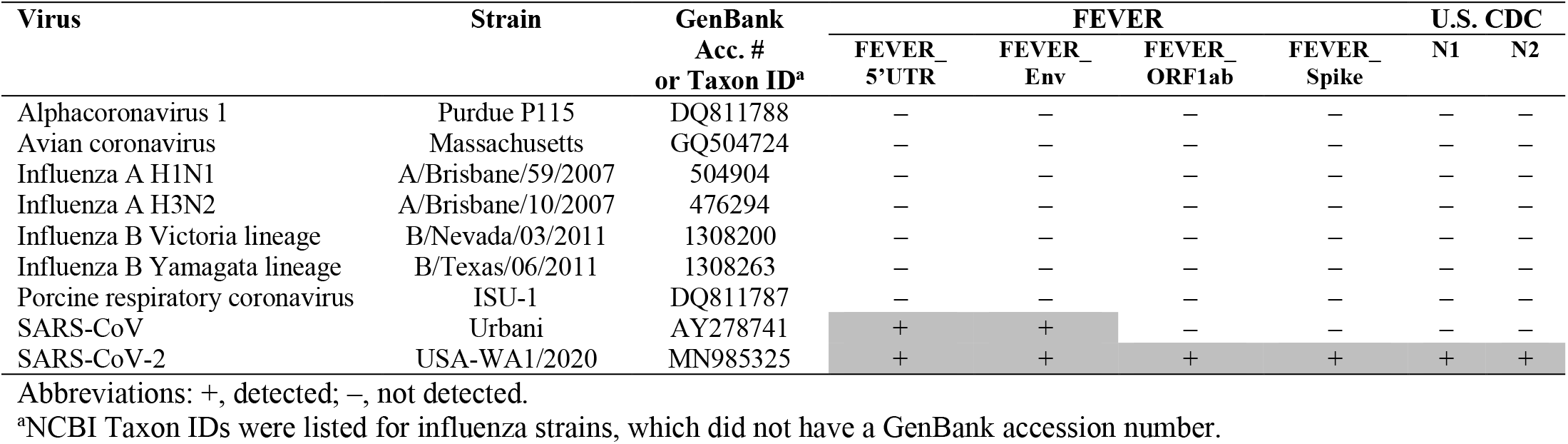
Analytical specificity of the FEVER and U.S. CDC assays.

### Analytical sensitivity in a NP swab matrix

The analytical sensitivity and reproducibility of each assay was tested using standard curves generated by three biological replicates of 10-fold serial dilutions of SARS-CoV-2 spiked into human NP swab samples. The standard curves for RT-PCR in NP swab samples had correlation coefficients (efficiency values) of 0.98 (100.9%) for FEVER_5’UTR, 0.99 (93.7%) for FEVER_Env, 0.99 (96.5%) for FEVER_ORF1ab, 0.99 (98.0%) for FEVER_Spike, 0.99 (103.2%) for N1, and 0.99 (99.1%) for N2, respectively (Fig. 1). We demonstrated that both FEVER and U.S. CDC assays reliably detected SARS-CoV-2 at 1 copy/µL spiked into an NP swab sample matrix.

**Figure 1.**
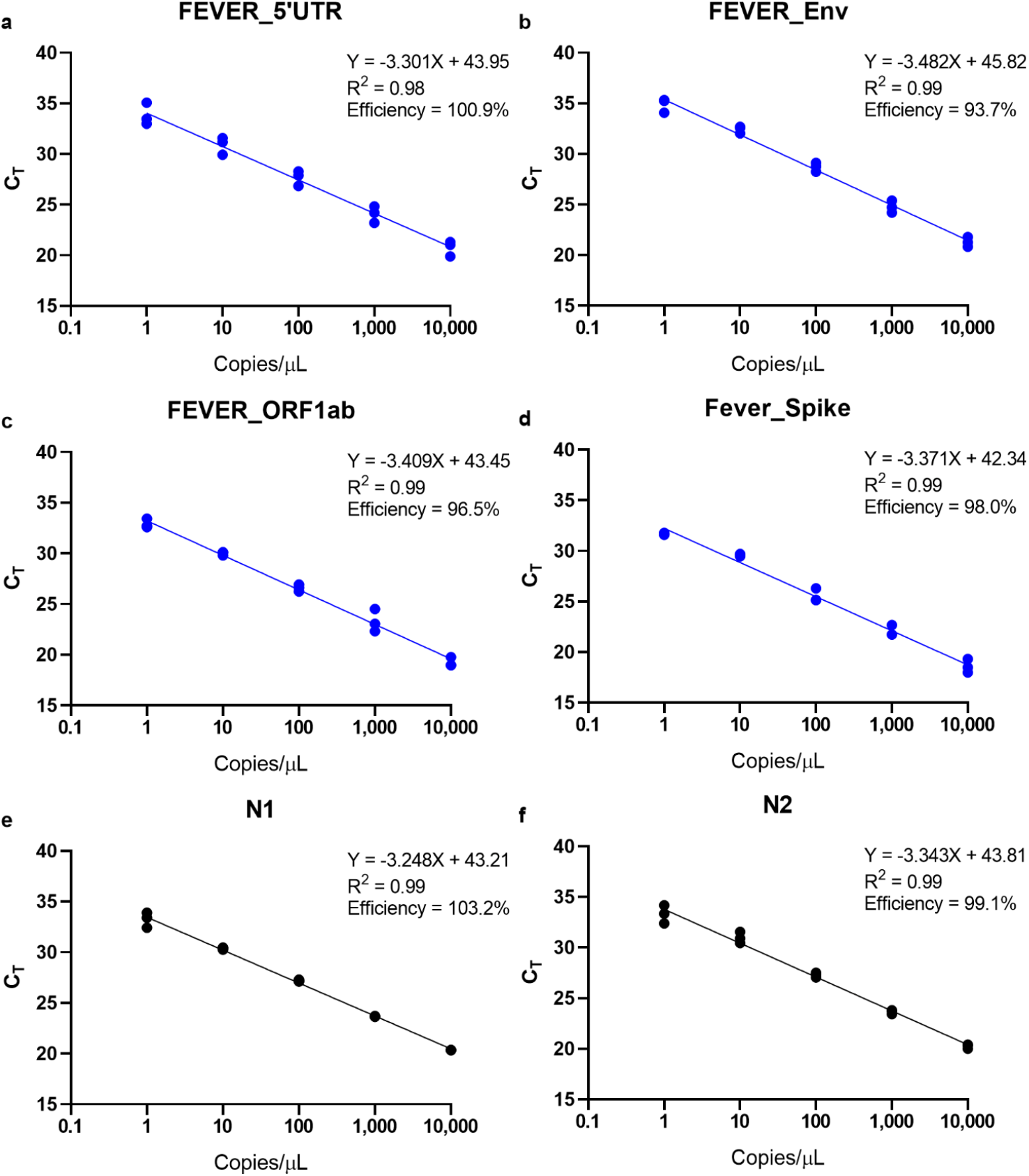
FEVER assays can detect 1 copy/µL SARS-CoV-2 spiked into a nasopharyngeal swab sample matrix. Inactivated SARS-CoV-2 strain USA-WA1/2020 was spiked into human nasopharyngeal swab samples (confirmed to be SARS-CoV-2 negative) and serial 10-fold diluted from 10^4^ to 1 copies/µL. Cycle threshold (C_T_) values were determined for FEVER assays (blue dots) (**a**) FEVER_5’UTR, (**b**) FEVER_Env, (**c**) FEVER_ORF1ab, and (**d**) FEVER_Spike and the (**a**) N1 and (**b**) N2 assay described by the U.S. CDC (black dots).

### Comparison of FEVER and U.S. CDC RT-PCR results for clinical NP swab samples

Nasopharyngeal (NP) swab sampling is the recommended sampling method by the World Health Organization (51) for nucleic acid amplification testing of SARS-CoV-2. Viral titers measured in NP swabs vary depending on several factors, including disease progression (52-54). However, NP swabs currently remain the reference sample type and support sensitive detection of SARS-CoV-2 (55). Nasopharyngeal swabs from 100 COVID-19 patients were collected in New Mexico, U.S. All assays were run under the same thermocycling conditions as per the U.S. CDC protocol (26). All samples tested positive for the human RNase P gene used as a control to ensure success of the RNA extraction ad associated sample collection methods (Table S1). Of the 100 NP swab samples, 66% (66/100) were considered positive according to the U.S. CDC assays and testing criteria (37), 22 were negative and 12 samples yielded inconclusive results (Table 4). Samples that initially produced an inconclusive result were re-tested and the 12 that remained inconclusive following the second test were removed from the analysis of agreement, sensitivity, and specificity. We compared the four FEVER assays to the results obtained with the U.S. CDC assays and found that all four FEVER assays displayed 100% specificity (no false positives were detected) and varying sensitivities ranging from 74.2% to 92.4% (Table 4, Table S1). However, the U.S. CDC assays were performed in a Clinical Laboratory Improvement Amendments (CLIA)-certified laboratory, while the FEVER assay was performed in a separate facility on a different thermocycler for research purposes only.

**Table 4.** FEVER assays compared to the U.S. CDC assays for detection of SARS-CoV-2 in 100 human patient nasopharyngeal swab samples.

| Result<sup>a</sup> | FEVER 5'UTR | FEVER Env | FEVER ORF1ab | FEVER Spike |
| --- | --- | --- | --- | --- |
| Positive | 61 | 56 | 56 | 49 |
| Negative | 39 | 44 | 44 | 51 |
| Agreement<sup>b</sup> | 94.3% (87.1-97.9%) | 87.5% (78.8-93.0%) | 88.6% (80.2-93.9%) | 80.7% (71.1-87.7%) |
| Sensitivity<sup>c</sup> | 92.4% (83.1-97.1%) | 83.3% (72.4-90.6%) | 84.8% (74.1-91.8%) | 74.2% (62.5-83.3%) |
| Specificity<sup>d</sup> | 100% (84.6-100%) | 100% (84.6-100%) | 100% (84.6-100%) | 100% (84.6-100%) |

<sup>a</sup> True values are as measured by the U.S. CDC assay criteria (37): 66 positive, 22 negative and 12 inconclusive. Samples initially recorded as inconclusive were re-tested and the 12 that remained inconclusive were removed from the analysis of agreement, sensitivity, and specificity.
<sup>b</sup>Agreement = no. of samples with identical results for an individual FEVER assay and U.S. CDC assay / 88
<sup>c</sup>Sensitivity = true positive / (true positive + false negative) with 95% confidence intervals
<sup>d</sup>Specificity = true negative / (true negative + false positive) with 95% confidence intervals
95% confidence intervals were calculated using the modified Wald method.
N/A, not applicable.

To directly compare the performance of the FEVER and U.S. CDC assays, 10 samples that contained sufficient remaining RNA were analyzed using both groups of assays on the same ABI StepOnePlus thermocycler in the same laboratory to avoid inter-instrument and inter-laboratory variability. The C_T_ values of the FEVER assays were plotted against those from the U.S. CDC assay (Fig. 2). In general, a broad range of C_T_ values from 13 to 39 was observed. Interestingly, one sample that originally tested positive for the U.S. CDC assay (N2) with a C_T_ value of 34 on the ABI 7500 Fast Dx, tested negative on the ABI StepOnePlus. There was a significant (*P* < 0.001), correlation between the C_T_ values of all four FEVER assays compared with each of the U.S. CDC N1 and N2 assays indicating that when run on the same instrument the FEVER and U.S. CDC assays yield highly similar C_T_values (Fig. 2). It is unfortunate that this finding occurred after most of the patient samples were exhausted and so not all could be re-tested using the same instrument. These results highlight the importance of using the same instrument when comparing C_T_values for assay validation.

**Figure 2.**
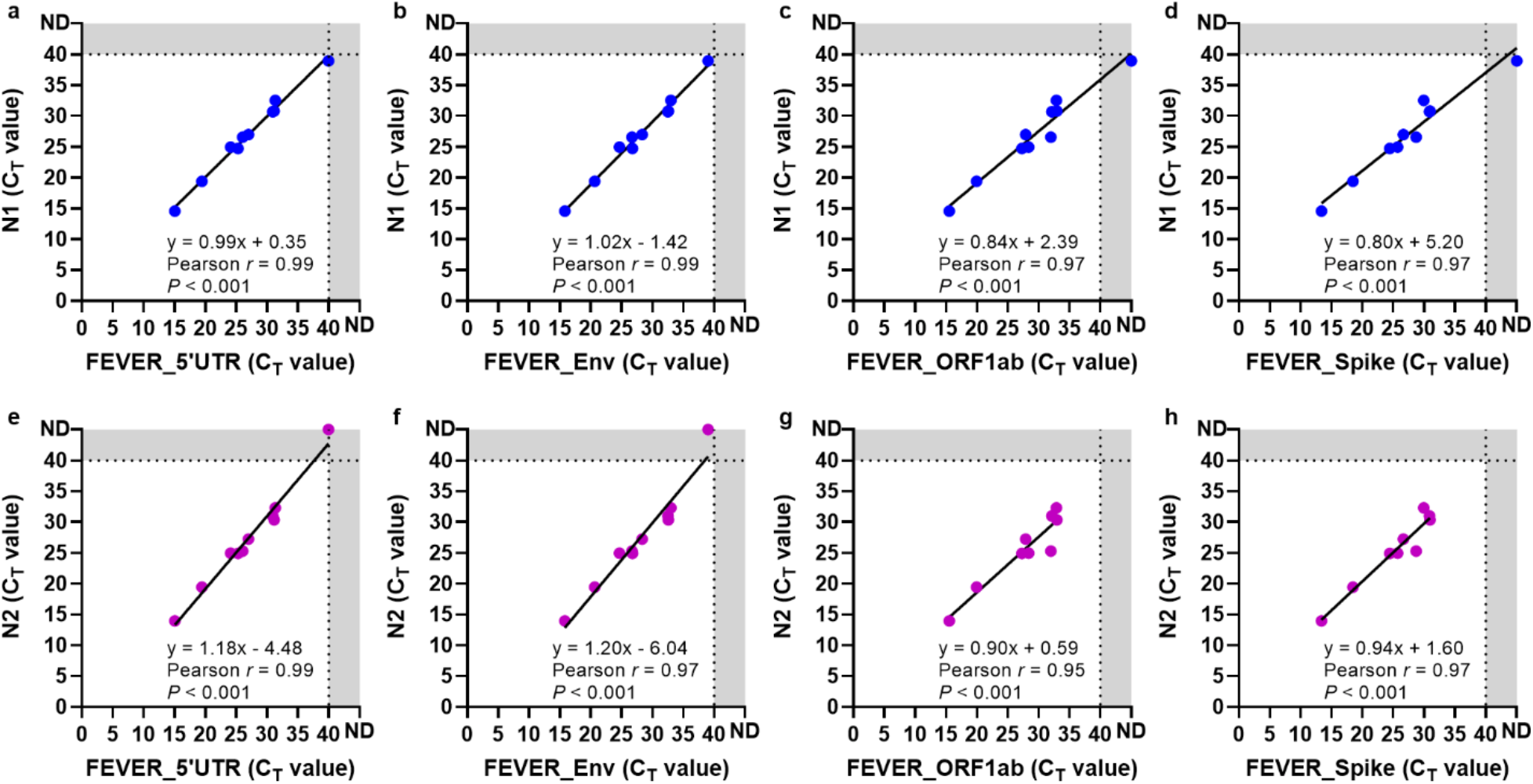
The FEVER and U.S. CDC assays show concordance in detecting SARS-CoV-2 in patient nasopharyngeal swab samples. The U.S. CDC N1 cycle threshold (C_T_) values were compared to (**a**) FEVER_5’UTR, (**b**) FEVER_Env, (**c**) FEVER_ORF1ab, and (**d**) FEVER_Spike RT-PCR assays C_T_values (blue dots). The U.S. CDC N2 C_T_ values were compared to (**e**) FEVER_5’UTR, (**f**) FEVER_Env, (**g**) FEVER_ORF1ab, and (**h**) FEVER_Spike RT-PCR assays C_T_ values (purple dots). One sample was removed for the N2 vs FEVER_ORF1ab and N2 vs FEVER_Spike comparisons because it was not detected by these assays. Each dot represents an individual sample. Dotted lines represent the limit of detection. ND, not detected.

### D614G mutation typing

High-throughput tracking of SARS-CoV-2 mutations in patient samples may be essential for monitoring the spread of vaccine escape mutants (42). Sequencing can identify novel variants but is too expensive and time-consuming for high-throughput screening of patient samples to track the spread and movement of these variants on a global scale (40). To facilitate a rapid and high-throughput method for monitoring the spread of SNPs of interest, we have incorporated a SNP Genotyping PCR assay. For this mutation typing test, we are detecting the A23403G (D614G amino acid change) mutation in the spike gene, which is associated with greater infectivity (40) (Table 1). Control RNA from isolate USA-WA1/2020 (BEI Resources, NR-52347) was used as a positive control, and results showed that USA-WA1/2020 tested positive for D614 (data not shown). Of the 59 SARS-CoV-2 positive patient samples (according to the FEVER assays), we determined the SNP at position 23,403 for 57 (96.6%) samples. The G23403 variant was found in 57 samples and 2 samples were undetermined with a C_T_ cutoff of 45 (Table S2). A fragment of the spike gene was amplified, and the A23403G SNP was confirmed in a subset of samples (n=17) by sequencing (Table S3). These results indicate that most (57/59) patient isolates contained the mutated G614 form of SARS-CoV-2 that became prevalent globally in April 2020 (40).

## DISCUSSION

Early detection of emerging pathogens is required to mitigate outbreaks and prevent pandemics (10). Assays used for diagnosing infections are often used for biosurveillance; however, due to their high specificity, they are not usually suited for broad-spectrum surveillance applications or capable of identifying pathogen mutations (43, 56). Therefore, the user must often decide between either obtaining diagnostics or biosurveillance information but not both. FEVER (Fast Evaluation of Viral Emerging Risks) is a computational approach designed to simultaneously facilitate broad-spectrum biosurveillance, pathogen-specific diagnostics, and SNP variant tracking for pandemic response to any viral pathogen of interest. In this manuscript, we demonstrate the application of FEVER to the ongoing COVID-19 pandemic as an initial proof-of-concept. As a proof of concept, we present FEVER assays to detect all sarbecoviruses for future biosurveillance, SARS-CoV-2 for strain-specific diagnostics, and the D614G spike variant SNP mutation associated with increased transmissibility.

Here, we used an RT-PCR format to evaluate the performance of the FEVER assays as compared to the U.S. CDC assays. The success of RT-PCR as a diagnostic approach relies heavily on the design quality of the primers and probes (57-59) as well as factors such as probe format, the ability to incorporate degenerate nucleotides, the number of target genomes that can be included in the design process to account for genetic diversity, and the ability to avoid cross-reactivity with near-neighbor genomes (60). In the case of SARS-CoV-2 diagnostics, it is also important that assays are able to differentiate infections caused by other common coronaviruses, in which case a false positive could result in unnecessary and costly mitigation procedures (61). For this reason, the U.S. CDC designed its N3 pan-*Sarbecovirus* assay; however, it was unfortunately removed from their assay in March of 2020 in order to expedite reagent manufacturing, consequently removing the biosurveillance component of the U.S. CDC COVID-19 assay (35, 37). This situation highlights the urgent need for a computational approach that can develop both biosurveillance and diagnostics assays that are screened for potential design issues that may potentially affect their performance and manufacturing.

Current probe design methods are also limited by over-representing single isolates in a sequence database as indicated by high coverage *in silico* but assay failure when novel variants emerge (31). In the case of SARS-CoV-2, the high number of false-negative results reported highlights the need for screening and updating assay designs as sequences become available (62-64), which has been a primary goal of https://cov.lanl.gov. Mutations in the probe or primer binding sites are particularly problematics since these can lead to false-negative results and infected individuals may continue to spread the disease (65). Pandemic preparedness requires a balanced approach combining targeted diagnostics assays designed to detect a specific outbreak strain with high-coverage biosurveillance assays that can account for pathogen mutation, and ideally this approach can be utilized to detect a variety of viral pathogens including those that are highly mutagenic. FEVER was designed to fill this void. In the case of SARS CoV-2, our assays were developed within a month of the U.S. CDC assays also using publicly available sequence data, but with a different computational approach in mind. The U.S. CDC designed highly specific diagnostics assays by pre-selecting a single genomic target, the viral nucleocapsid gene, aligned a reference SARS-CoV-2 sequence (GenBank accession no. MN908947), and selected probe/primer sets with low similarity to other coronavirus sequences in GenBank (37). This method has yielded two SARS-CoV-2 specific assays that have contributed to combatting the current pandemic, however it is unclear how useful these assays will be in detecting the next coronavirus outbreak.

FEVER takes a more comprehensive approach to design robust multi-purpose assays to detect circulating viral strains and track their mutations but also to detect entire classes of viral pathogens in order to monitor emerging threats. The FEVER algorithm scans the entire viral genome to identify conserved regions and identifies the lowest possible number of probes necessary to cover all sequences scanned. The diagnostics probe FEVER_ORF1ab displayed the highest coverage of SARS-CoV-2 sequences in this study with a predicted recall of 99.7%. It was predicted to detect all but 1,454 SARS-CoV-2 sequences out of 525,708 deposited in GISAID as of February 15, 2021. In comparison, the U.S. CDC N2 probe displayed 99.17% coverage and was predicted to fail in 4,353 sequences, which is nearly a 3-fold increase in false-negative results.

In addition to optimized sequence analysis, FEVER incorporates physical constraints set by the user to include GC-content, hairpin propensity, sequence length, and even mismatch tolerance. This ensures that probes selected for testing are thermodynamically compatible for increased assay performance as well as future multiplexing. It is critical that assay sensitivity is optimized as a 10-fold increase in the limit of detection for SARS-CoV-2 has shown to result in an increase in the false-negative rate by 13% (66). Here, the limit of detection for both the FEVER and U.S. CDC assays in NP swab samples was 1 copy/µL which matches previously published findings (26). Additionally, FEVER assay design features can be used to ensure that probes and primers are suitable for large-scale manufacturing and can be tailored to a variety of biosensing platforms, including but not limited to RT-PCR. There is a clear need for the development of rapid and accurate point-of-care diagnostic tools to detect viral infections (67), and a variety of nucleic acid-based viral biosensors are in development for this purpose (43).

FEVER also includes a method for rapid mutation typing to track viral variants of public health importance. SARS-CoV-2 has proofreading mechanisms (68) and therefore relatively lower genetic diversity than other respiratory viruses such as influenza (69). For SARS-CoV-2, the predicted evolutionary rate is approximately 8×10^−4^ nucleotide substitution per site per year (69). The spike gene is under immune selective pressure as neutralizing antibodies against the protein product can inhibit viral entry into host cells (40, 70). SARS-CoV-2 acquired the D614G mutation in the spike protein early in the pandemic, and this mutation confers greater infectivity (71-73). Previously, the detection of this mutation has been based solely on viral sequencing, which is time-consuming and cost-prohibitive in many resource limited regions of the world (40, 61, 74, 75). FEVER incorporates a rapid PCR-based method for identifying the D614G mutation directly in patient samples. Similarly, mutation typing probes and primers can be designed in the future to track other variants of interest including the U.K. (B.1.1.7), South Africa (B.1.351), and Brazil (P.1) variants (41, 76, 77) to provide insight and real-time biosurveillance data on the circulation of potential vaccine escape variants.

In summary, the FEVER computational approach provides a novel tool to combat emerging viral pathogens. FEVER can be used for simultaneous biosurveillance of viruses, diagnostics of outbreak or pandemic isolates, and mutation typing to rapidly track the spread of variants impacting public health. In addition to PCR, FEVER can generate probes that are compatible with a wide variety of nucleic acid sensing techniques which may be more suitable to point-of-care testing than RT-PCR (43). Here we show that the FEVER assays are comparable to those designed by the U.S. CDC for the detection of SARS-CoV-2 while simultaneously developing pan-*Sarbecovirus* assays for future biosurveillance applications. Additionally, FEVER mutation typing assays provide a rapid means for monitoring the spread of potential vaccine escape variants that may significantly impact a population. Future work will focus on applying FEVER towards the development of assays for the detection of other upper respiratory pathogens including influenza as well as testing FEVER probes in ultra-sensitive amplification-free nucleic acid biosensing platforms. FEVER provides a holistic computational approach to combine biosurveillance and diagnostics in order to better combat emerging viral pathogens.

## SUPPORTING INFORMATION

**Figure S1. Visualization of SARS-CoV-2 variants in the amplified regions of FEVER and U. S. CDC RT-PCR assays**. Near-complete SARS-CoV-2 sequences (n = 299,404) were assessed from GISAID.org. The amplified regions of the FEVER assays (a) FEVER_5’UTR, (b) FEVER_Env, (c) FEVER_ORF1ab, and (d) FEVER_Spike and the U.S. CDC assays (e) N1 and (f) N2 were analyzed for SARS-CoV-2 variants. In each plot, no variants (N/A) were indicated by white and genomic positions containing variants were indicated by colored dots (green, blue, yellow or red). Plots were made using the Variant Visualizer that is under development and will be available at: https://cov.lanl.gov.

**Table S1. Raw C**_**T**_ **values of FEVER and U.S. CDC assays run on 100 human patient nasopharyngeal swab samples.**

**Table S2. Characterization of the D614G mutation among 59 SARS-CoV-2 positive patient nasopharyngeal swab samples.**

**Table S3. Sequence confirmation of the D614G mutation among 17 selected patient nasopharyngeal swab samples.**

## ACKNOWLEDGMENTS

This research was funded by Los Alamos National Laboratory Exploratory Research and Development grant number 20190392ER (to J.K.S and K.Y.) and Los Alamos National Laboratory Technology Evaluation and Demonstration Funds. The authors would like to thank the Presbyterian Clinical Research team (Brigitte Holder, Susan Salazar, and Maria Vahtel) for their contributions to this study. We are also grateful to the patients who volunteered to take part in this study.

