## Supplemental Material for "Fast Evaluation of Viral Emerging Risks (FEVER): A computational tool for biosurveillance, diagnostics, and mutation typing of emerging viral pathogens"

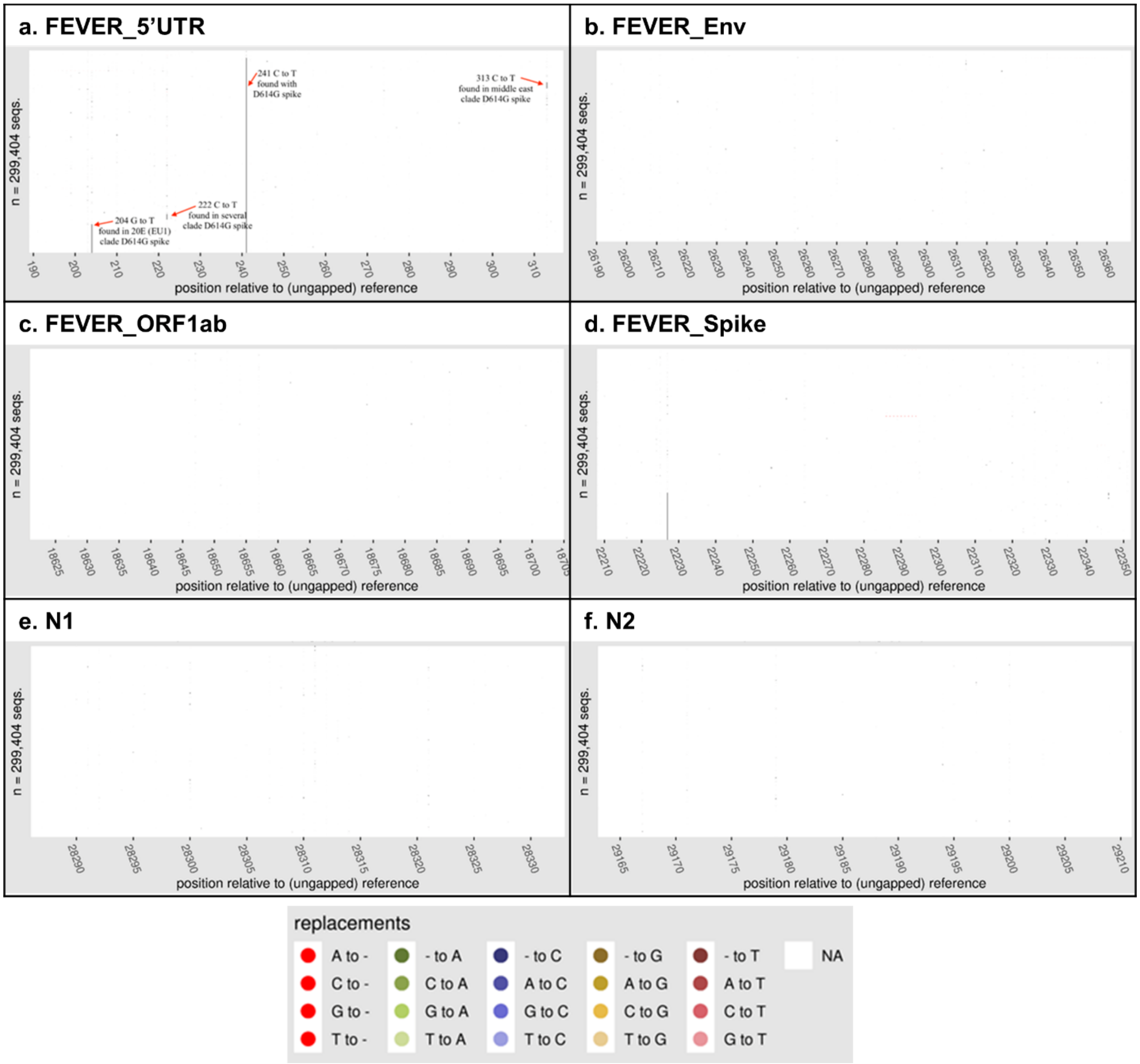


**Figure S1. Visualization of SARS-CoV-2 variants in the amplified regions of FEVER and U. S. CDC RT-PCR assays.** Near-complete SARS-CoV-2 sequences (n = 299,404) were assessed from GISAID.org. The amplified regions of the FEVER assays (a) FEVER_5'UTR, (b) FEVER_Env, (c) FEVER_ORF1ab, and (d) FEVER_Spike and the U.S. CDC assays (e) N1 and (f) N2 were analyzed for SARS-CoV-2 variants. In each plot, no variants (N/A) were indicated by white and genomic positions containing variants were indicated by colored dots (green, blue, yellow or red). Plots were made using the Variant Visualizer that is under development and will be available at: https://cov.lanl.gov.

**Table S1. Raw CT values of FEVER and U.S. CDC assays run on 100 human patient nasopharyngeal swab samples.**

| | $C_T$ Value | | | | | | | Result |
|---|---|---|---|---|---|---|---|---|
| | pan-*Sarbecovirus* | | SARS-CoV-2 specific | | | | Human Control | |
| **Sample ID** | **FEVER_5'UTR** | **FEVER_Env** | **FEVER_ORF1ab** | **FEVER_Spike** | **U.S. CDC N1** | **U.S. CDC N2** | **RNase P** | **U.S. CDC** |
| 1 | 31.32 | 32.98 | 32.87 | 29.96 | 26.07 | 26.48 | 26.97 | Positive |
| 2 | 25.29 | 26.76 | 27.27 | 24.44 | 19.46 | 20.11 | 25.91 | Positive |
| 3 | 15.07 | 15.79 | 15.50 | 13.35 | 10.54 | 11.47 | 28.48 | Positive |
| 4 | Not detected | Not detected | Not detected | Not detected | Not detected | 39.85 | 25.62 | Inconclusive |
| 5 | Not detected | Not detected | Not detected | Not detected | Not detected | Not detected | 31.11 | Negative |
| 6 | 26.99 | 28.32 | 27.89 | 26.64 | 21.59 | 22.26 | 29.80 | Positive |
| 7 | 31.14 | 32.57 | 32.93 | 30.95 | 25.59 | 27.46 | 30.73 | Positive |
| 8 | 30.90 | 32.49 | 32.11 | 30.87 | 26.30 | 28.02 | 30.97 | Positive |
| 9 | 19.43 | 20.65 | 19.93 | 18.45 | 14.77 | 16.07 | 26.05 | Positive |
| 10 | Not detected | Not detected | Not detected | Not detected | Not detected | Not detected | 31.40 | Negative |
| 11 | 40.97 | Not detected | Not detected | Not detected | 34.81 | 42.52 | 28.76 | Inconclusive |
| 12 | Not detected | Not detected | Not detected | Not detected | Not detected | Not detected | 32.72 | Negative |
| 13 | Not detected | Not detected | Not detected | Not detected | Not detected | Not detected | 34.49 | Negative |
| 14 | Not detected | Not detected | Not detected | Not detected | Not detected | Not detected | 31.25 | Negative |
| 15 | 21.09 | 20.60 | 23.57 | 22.28 | 16.35 | 16.96 | 24.60 | Positive |
| 16 | 24.08 | 24.66 | 28.38 | 25.69 | 19.88 | 21.10 | 31.82 | Positive |
| 17 | 26.05 | 26.68 | 31.97 | 28.69 | 20.49 | 21.08 | 31.28 | Positive |
| 18 | 37.16 | 38.99 | Not detected | 32.98 | 32.75 | 33.27 | 31.26 | Positive |
| 19 | 25.05 | 25.46 | 28.56 | 27.35 | 20.17 | 20.50 | 28.61 | Positive |
| 20 | 26.13 | 27.35 | 30.98 | 30.93 | 20.70 | 21.38 | 25.95 | Positive |
| 21 | 32.28 | 31.66 | 33.93 | 27.74 | 27.10 | 27.74 | 27.40 | Positive |
| 22 | 24.01 | Not detected | Not detected | 21.47 | 17.29 | 19.17 | 23.06 | Positive |
| 23 | 25.54 | 25.12 | 26.60 | Not detected | 20.21 | 21.05 | 26.87 | Positive |
| 24 | 34.16 | 33.56 | 33.58 | 31.74 | 29.77 | 30.65 | 27.90 | Positive |
| 25 | 27.91 | 28.64 | 26.39 | Not detected | 22.27 | 23.03 | 25.18 | Positive |
| 26 | Not detected | Not detected | Not detected | Not detected | Not detected | Not detected | 29.48 | Negative |
| 27 | Not detected | Not detected | Not detected | Not detected | Not detected | Not detected | 25.26 | Negative |
| 28 | 21.14 | 35.98 | 18.90 | Not detected | 16.75 | 17.30 | 27.09 | Positive |
| 29 | 28.64 | 31.18 | 28.65 | Not detected | 23.54 | 24.06 | 23.98 | Positive |
| 30 | 28.26 | 34.59 | 26.72 | 24.67 | 22.59 | 23.49 | 23.46 | Positive |
| 31 | 28.71 | 28.75 | 27.56 | 24.93 | 24.39 | 24.66 | 25.23 | Positive |
| 32 | Not detected | Not detected | Not detected | Not detected | Not detected | Not detected | 28.24 | Negative |
| 33 | 13.43 | 11.96 | 12.84 | 11.02 | 13.66 | 13.82 | 18.64 | Positive |
| 34 | Not detected | Not detected | Not detected | Not detected | Not detected | Not detected | 30.72 | Negative |
| 35 | 33.53 | 33.35 | 36.64 | 30.95 | 28.46 | 29.61 | 23.98 | Positive |
| 36 | 18.01 | 16.77 | 16.37 | 15.70 | 15.37 | 15.79 | 26.92 | Positive |
| 37 | 26.02 | 25.16 | 24.98 | 24.36 | 22.24 | 23.06 | 24.59 | Positive |
| 38 | 15.00 | 13.71 | 13.42 | 12.56 | 14.19 | 14.22 | 28.85 | Positive |
| 39 | 16.99 | 16.29 | 16.03 | 14.97 | 14.78 | 15.05 | 25.54 | Positive |
| 40 | 28.95 | 28.23 | 27.78 | 30.90 | 25.27 | 25.66 | 28.84 | Positive |
| 41 | Not detected | Not detected | Not detected | Not detected | Not detected | 37.31 | 30.43 | Inconclusive |
| 42 | 28.84 | 41.00 | 29.00 | 29.72 | 24.79 | 25.33 | 26.33 | Positive |
| 43 | 32.77 | 32.74 | 33.90 | 31.89 | 29.00 | 29.38 | 28.32 | Positive |
| 44 | 29.28 | 30.10 | 30.75 | 28.89 | 25.82 | 26.24 | 29.31 | Positive |
| 45 | Not detected | Not detected | Not detected | Not detected | Not detected | Not detected | 27.78 | Negative |
| 46 | 32.45 | 35.69 | 32.72 | Not detected | 29.20 | 29.37 | 31.97 | Positive |
| 47 | Not detected | Not detected | Not detected | Not detected | Not detected | Not detected | 28.15 | Negative |
| 48 | Not detected | Not detected | Not detected | Not detected | 35.81 | Not detected | 25.32 | Inconclusive |
| 49 | 37.00 | Not detected | 34.34 | Not detected | 30.88 | 31.64 | 26.97 | Positive |
| 50 | 30.24 | Not detected | 33.72 | 30.93 | 28.24 | 29.43 | 24.57 | Positive |
| 51 | Not detected | 42.03 | Not detected | Not detected | 32.45 | 34.11 | 28.98 | Positive |
| 52 | 30.65 | 37.20 | 33.75 | 31.28 | 29.25 | 30.14 | 25.82 | Positive |
| 53 | Not detected | Not detected | Not detected | Not detected | Not detected | Not detected | 23.32 | Negative |
| 54 | 29.52 | 40.98 | 31.72 | 29.99 | 28.58 | 29.57 | 26.81 | Positive |
| 55 | Not detected | 35.96 | 34.87 | Not detected | 34.16 | 35.09 | 30.42 | Positive |
| 56 | 40.54 | 37.75 | Not detected | Not detected | 33.26 | 35.64 | 34.48 | Positive |
| 57 | 35.57 | 38.20 | 32.89 | Not detected | 32.25 | 32.87 | 32.41 | Positive |
| 58 | 25.61 | 26.92 | 29.69 | 27.59 | 19.89 | 21.22 | 24.34 | Positive |
| 59 | 29.37 | 30.15 | 31.96 | 29.90 | 24.95 | 26.53 | 26.02 | Positive |
| 60 | 20.01 | 20.19 | 22.14 | 20.64 | 15.43 | 16.54 | 29.19 | Positive |
| 61 | Not detected | Not detected | Not detected | Not detected | Not detected | Not detected | 30.48 | Negative |
| 62 | 29.47 | 32.99 | Not detected | Not detected | 24.67 | 25.96 | 26.16 | Positive |
| 63 | 34.96 | 33.67 | 30.85 | Not detected | 29.80 | 30.64 | 29.15 | Positive |
| 64 | Not detected | Not detected | 34.86 | Not detected | 33.93 | 34.70 | 30.38 | Positive |
| 65 | Not detected | Not detected | Not detected | Not detected | Not detected | 37.69 | 32.29 | Inconclusive |
| 66 | 32.80 | Not detected | Not detected | 30.38 | 26.07 | 28.08 | 31.42 | Positive |
| 67 | 20.13 | 24.57 | 25.46 | 21.63 | 15.22 | 16.38 | 30.91 | Positive |
| 68 | Not detected | Not detected | Not detected | Not detected | Not detected | Not detected | 28.84 | Negative |

| Sample ID | FEVER_5'UTR | FEVER_Env | FEVER_ORF1ab | FEVER_Spike | U.S. CDC N1 | U.S. CDC N2 | RNase P | U.S. CDC |
|---|---|---|---|---|---|---|---|---|
| 69 | Not detected | Not detected | Not detected | Not detected | Not detected | Not detected | 26.98 | Negative |
| 70 | 40.97 | Not detected | Not detected | Not detected | Not detected | 36.30 | 32.80 | Inconclusive |
| 71 | Not detected | Not detected | Not detected | Not detected | Not detected | Not detected | 29.72 | Negative |
| 72 | Not detected | Not detected | Not detected | Not detected | 35.28 | Not detected | 26.26 | Inconclusive |
| 73 | 30.98 | 30.26 | 32.99 | 31.87 | 26.54 | 28.06 | 30.91 | Positive |
| 74 | Not detected | Not detected | Not detected | Not detected | Not detected | 36.42 | 29.59 | Inconclusive |
| 75 | Not detected | Not detected | Not detected | Not detected | Not detected | Not detected | 27.10 | Negative |
| 76 | 19.77 | 18.40 | 22.39 | 22.51 | 15.26 | 16.16 | 24.85 | Positive |
| 77 | 27.03 | 26.03 | 28.92 | 28.19 | 22.45 | 24.11 | 29.37 | Positive |
| 78 | 24.80 | 24.19 | 28.73 | 26.06 | 20.62 | 21.72 | 25.73 | Positive |
| 79 | 38.80 | 38.98 | Not detected | Not detected | 32.47 | 35.74 | 23.71 | Positive |
| 80 | 26.30 | 27.02 | 29.09 | 27.67 | 21.10 | 22.39 | 29.75 | Positive |
| 81 | Not detected | Not detected | Not detected | Not detected | Not detected | Not detected | 30.68 | Negative |
| 82 | Not detected | Not detected | Not detected | Not detected | Not detected | 35.44 | 30.98 | Inconclusive |
| 83 | 35.90 | 34.08 | 33.88 | 32.98 | 27.01 | 28.83 | 24.94 | Positive |
| 84 | 39.10 | 38.92 | Not detected | Not detected | 35.58 | 37.83 | 25.81 | Positive |
| 85 | Not detected | Not detected | Not detected | Not detected | 34.44 | 35.69 | 30.30 | Positive |
| 86 | Not detected | Not detected | Not detected | Not detected | Not detected | 36.49 | 30.49 | Inconclusive |
| 87 | Not detected | Not detected | Not detected | Not detected | 33.90 | 41.12 | 31.98 | Inconclusive |
| 88 | 31.64 | Not detected | 31.70 | 29.36 | 26.53 | 28.26 | 28.98 | Positive |
| 89 | 24.80 | 22.34 | 25.85 | 22.21 | 21.18 | 22.85 | 33.10 | Positive |
| 90 | Not detected | Not detected | Not detected | Not detected | Not detected | Not detected | 29.31 | Negative |
| 91 | 27.62 | 24.73 | 26.14 | 24.78 | 22.15 | 23.84 | 30.30 | Positive |
| 92 | 15.32 | 13.00 | 14.23 | 12.91 | 14.21 | 14.70 | 31.69 | Positive |
| 93 | 38.97 | Not detected | Not detected | Not detected | 30.07 | 32.36 | 25.91 | Positive |
| 94 | Not detected | Not detected | Not detected | Not detected | Not detected | Not detected | 28.43 | Negative |
| 95 | 32.15 | 31.45 | 33.88 | 30.95 | 28.11 | 29.93 | 23.93 | Positive |
| 96 | Not detected | 37.01 | Not detected | Not detected | 37.62 | Not detected | 19.39 | Inconclusive |
| 97 | 24.19 | 22.58 | 25.02 | 23.42 | 20.54 | 22.51 | 28.40 | Positive |
| 98 | Not detected | Not detected | Not detected | Not detected | Not detected | Not detected | 29.97 | Negative |
| 99 | 26.08 | 23.60 | 26.39 | 31.93 | 22.31 | 24.52 | 31.22 | Positive |
| 100 | Not detected | Not detected | Not detected | Not detected | Not detected | Not detected | 32.46 | Negative |

**Table S2. Characterization of the D614G mutation among 59 SARS-CoV-2 positive nasopharyngeal swab samples.**

| Sample # | SARS-CoV-2 FEVER result | G614 $C_T$ | D614 $C_T$ | SNP call |
|---|---|---|---|---|
| 1 | Positive | 33.54 | 36.09 | G614 |
| 2 | Positive | 24.55 | 26.92 | G614 |
| 3 | Positive | 12.95 | 15.27 | G614 |
| 6 | Positive | 25.93 | 28.67 | G614 |
| 7 | Positive | 31.81 | 34.16 | G614 |
| 8 | Positive | 31.99 | 34.62 | G614 |
| 9 | Positive | 18.69 | 21.64 | G614 |
| 15 | Positive | 23.77 | 26.64 | G614 |
| 16 | Positive | 22.41 | 24.93 | G614 |
| 17 | Positive | 24.74 | 27.51 | G614 |
| 18 | Positive | 38.68 | 41.46 | G614 |
| 19 | Positive | 23.21 | 25.71 | G614 |
| 20 | Positive | 25.97 | 28.83 | G614 |
| 21 | Positive | 30.69 | 33.27 | G614 |
| 22 | Positive | 27.56 | 30.31 | G614 |
| 23 | Positive | 25.20 | 27.71 | G614 |
| 24 | Positive | 34.50 | 36.95 | G614 |
| 25 | Positive | 25.91 | 28.58 | G614 |
| 28 | Positive | 21.64 | 24.38 | G614 |
| 29 | Positive | 29.69 | 32.41 | G614 |
| 30 | Positive | 37.62 | 40.72 | G614 |
| 31 | Positive | 30.84 | 33.54 | G614 |
| 33 | Positive | 14.38 | 17.16 | G614 |
| 35 | Positive | 34.30 | 36.78 | G614 |
| 36 | Positive | 19.00 | 21.86 | G614 |
| 37 | Positive | 27.32 | 29.89 | G614 |
| 38 | Positive | 15.96 | 18.64 | G614 |
| 39 | Positive | 17.52 | 20.23 | G614 |
| 40 | Positive | 32.84 | 35.60 | G614 |
| 42 | Positive | 29.46 | 32.00 | G614 |
| 43 | Positive | 34.50 | 37.24 | G614 |
| 44 | Positive | 29.90 | 32.76 | G614 |
| 46 | Positive | 31.98 | 34.90 | G614 |
| 49 | Positive | 35.34 | 38.11 | G614 |
| 50 | Positive | 33.47 | 36.36 | G614 |
| 52 | Positive | 35.60 | 38.36 | G614 |
| 54 | Positive | 33.62 | 36.43 | G614 |
| 55 | Positive | 39.91 | 44.01 | G614 |
| 57 | Positive | 37.39 | 41.02 | G614 |
| 58 | Positive | 27.05 | 30.97 | G614 |
| 59 | Positive | 32.81 | 36.61 | G614 |
| 60 | Positive | 21.72 | 25.52 | G614 |
| 62 | Positive | 37.77 | 41.58 | G614 |
| 63 | Positive | 34.80 | 38.69 | G614 |
| 66 | Positive | 29.46 | 33.33 | G614 |
| 67 | Positive | 19.23 | 23.00 | G614 |
| 73 | Positive | 33.84 | 36.52 | G614 |
| 76 | Positive | 19.34 | 21.92 | G614 |
| 77 | Positive | 37.29 | 40.39 | G614 |
| 78 | Positive | 24.74 | 27.25 | G614 |

| | | | | |
|---|---|---|---|---|
| 80 | Positive | 25.70 | 28.39 | G614 |
| 83 | Positive | 35.48 | 38.16 | G614 |
| 88 | Positive | 32.01 | 34.85 | G614 |
| 89 | Positive | 25.03 | 27.70 | G614 |
| 91 | Positive | 28.52 | 31.25 | G614 |
| 92 | Positive | 15.59 | 18.35 | G614 |
| 95 | Positive | 33.65 | 36.51 | G614 |
| 97 | Positive | Not detected | Not detected | Not detected |
| 99 | Positive | Not detected | Not detected | Not detected |

Abbreviations: $C_T$, cycle threshold; SNP, single-nucleotide polymorphism.

**Table S3. Sequence confirmation of the D614G mutation among 17 nasopharyngeal swab samples.**

| Sample | Sequence (23,353-23,452) |
|---|---|
| MN985325 (reference) | TATAACACCAGGAACAAATACTTCTAACCAGGTTGCTGTTCTTTATCAGG**A**TGTTAACTGCACAGAAGTCCCTGTTGCTATTCATGCAGATCAACTTACT |
| 1 | TATAACACCAGGAACAAATACTTCTAACCAGGTTGCTGTTCTTTATCAGG**G**TGTTAACTGCACAGAAGTCCCTGTTGCTATTCATGCAGATCAACTTACT |
| 2 | TATAACACCAGGAACAAATACTTCTAACCAGGTTGCTGTTCTTTATCAGG**G**TGTTAACTGCACAGAAGTCCCTGTTGCTATTCATGCAGATCAACTTACT |
| 3 | TATAACACCAGGAACAAATACTTCTAACCAGGTTGCTGTTCTTTATCAGG**G**TGTTAACTGCACAGAAGTCCCTGTTGCTATTCATGCAGATCAACTTACT |
| 6 | TATAACACCAGGAACAAATACTTCTAACCAGGTTGCTGTTCTTTATCAGG**G**TGTTAACTGCACAGAAGTCCCTGTTGCTATTCATGCAGATCAACTTACT |
| 8 | TATAACACCAGGAACAAATACTTCTAACCAGGTTGCTGTTCTTTATCAGG**G**TGTTAACTGCACAGAAGTCCCTGTTGCTATTCATGCAGATCAACTTACT |
| 9 | TATAACACCAGGAACAAATACTTCTAACCAGGTTGCTGTTCTTTATCAGG**G**TGTTAACTGCACAGAAGTCCCTGTTGCTATTCATGCAGATCAACTTACT |
| 16 | TATAACACCAGGAACAAATACTTCTAACCAGGTTGCTGTTCTTTATCAGG**G**TGTTAACTGCACAGAAGTCCCTGTTGCTATTCATGCAGATCAACTTACT |
| 17 | TATAACACCAGGAACAAATACTTCTAACCAGGTTGCTGTTCTTTATCAGG**G**TGTTAACTGCACAGAAGTCCCTGTTGCTATTCATGCAGATCAACTTACT |
| 18 | TATAACACCAGGAACAAATACTTCTAACCAGGTTGCTGTTCTTTATCAGG**G**TGTTAACTGCACAGAAGTCCCTGTTGCTATTCATGCAGATCAACTTACT |
| 21 | TATAACACCAGGAACAAATACTTCTAACCAGGTTGCTGTTCTTTATCAGG**G**TGTTAACTGCACAGAAGTCCCTGTTGCTATTCATGCAGATCAACTTACT |
| 22 | TATAACACCAGGAACAAATACTTCTAATCAGGTTGCTGTTCTTTATCAGG**G**TGTTAACTGCACAGAAGTCCCTGTTGCTATTCATGCAGATCAACTTACT |
| 30 | TATAACACCAGGAACAAATACTTCTAACCAGGTTGCTGTTCTTTATCAGG**G**TGTTAACTGCACAGAAGTCCCTGTTGCTATTCATGCAGATCAACTTACT |
| 52 | TATAACACCAGGAACAAATACTTCTAACCAGGTTGCTGTTCTTTATCAGG**G**TGTTAACTGCACAGAAGTCCCTGTTGCTATTCATGCAGATCAACTTACT |
| 59 | TATAACACCAGGAACAAATACTTCTAACCAGGTTGCTGTTCTTTATCAGG**G**TGTTAACTGCACAGAAGTCCCTGTTGCTATTCATGCAGATCAACTTACT |
| 73 | TATAACACCAGGAACAAATACTTCTAACCAGGTTGCTGTTCTTTATCAGG**G**TGTTAACTGCACAGAAGTCCCTGTTGCTATTCATGCAGATCAACTTACT |
| 78 | TATAACACCAGGAACAAATACTTCTAACCAGGTTGCTGTTCTTTATCAGG**G**TGTTAACTGCACAGAAGTCCCTGTTGCTATTCATGCAGATCAACTTACT |
| 92 | TATAACACCAGGAACAAATACTTCTAACCAGGTTGCTGTTCTTTATCAGG**G**TGTTAACTGCACAGAAGTCCCTGTTGCTATTCATGCAGATCAACTTACT |

The spike gene from nucleotide position 23,353 to 23,452 (relative to reference sequence USA-WA1/2020 accession number MN985325) is displayed with A23403G (D614G amino acid change) mutation in red.
